# Factors affecting zero-waste behaviors: Focusing on the health effects of microplastics

**DOI:** 10.1101/2022.03.04.22271923

**Authors:** Eun-Hi Choi, Hyunjin Lee, Mi-Jung Kang, Inwoo Nam, Hui-Kyeong Moon, Ji-Won Sung, Jae-Yun Eu, Hae-Bin Lee

**Affiliations:** College of Nursing, Eulji University, Uijeongbu-si, Gyeonggi-do, Republic of Korea

**Author notes:** Corresponding author (HL); (MJK). These authors contributed equally to this work.

**Keywords:** zero-waste, health, microplastics

## Abstract

Microplastics harm human health. Therefore, the present study assessed the knowledge and attitude of university students towards reducing microplastic use and examined their zero-waste behaviors. Our results lay the foundation for program development aimed at promoting zero-waste activities. The study was conducted from August 20, 2021, to September 10, 2021, including students at a university in G metropolitan city. Questions were developed to verify how the use of disposables and the knowledge, attitude, and behaviors related to zero-waste were affected after the COVID-19 pandemic. A survey was conducted with 197 students, and the data of 196 students were analyzed. Family type (β=0.146, p=0.042) and usage of disposables (β=0.158, p=0.049) were the factors affecting zero-waste behavior in Model 1. In Model 2, which included the subcategory of zero-waste knowledge, the health effects of microplastics (β=0.197, p=0.008) and environmental preservation (β=0.236, p=0.001) were significant factors. In Model 3, which included the subcategory of zero-waste attitude, the health effects of microplastics (β=0.149, p=0.016), use of eco-friendly products (β=0.342, p<0.001), and environmental preservation (β=0.317, p<.001) were significant factors. Therefore, additional studies and education on the health effects of microplastics are warranted, and suitable alternatives for disposables must be developed.

## Introduction

During the COVID-19 pandemic [1], the World Health Organization (WHO) recommended wearing masks, maintaining social distance, and avoiding gatherings [2]. To maintain social distancing, non-face-to-face classes using online platforms were started, and students attended classes from homes or dorm rooms without attending colleges. Consequently, they ended up spending more time in their living spaces [3] and frequently used home meal replacement or delivery food [4].

Given these changes in lifestyle during the pandemic, the demand for plastic products increased, producing a severe impact on the environment. Plastic waste management was already considered a major environmental issue before the COVID-19 pandemic [5-6]. After the outbreak, the demand for plastic disposables, such as convenience and delivery food containers, has dramatically increased [7], leading serious environmental issues. In particular, the annual usage of plastic per person in Korea is 132.7 kg, which is the highest in the world [8]. However, the current waste management system is not sufficiently effective to manage existing plastic waste [9], and the rapid increase in the amount of plastic waste due to COVID-19 is expected to lead to a bigger problem.

Plastic waste contains many harmful substances, with microplastic being the most hazardous material [10]. In previous studies, microplastics were detected in marine organisms, and harmful effects of microplastics in various aquatic organisms have been reported [11-12]. In addition, microplastics were detected in table salt [13], drinking water [14], and air [15], indicating that human exposure to microplastics is inevitable. Recent studies have reported the association of microplastics with the development of various diseases, including cancer [16-18].

These concerns regarding plastic waste led to the creation of a new concept of zero waste. Its definition differs depending on the purpose of the activity and position of the activity subject [19-21]. In 2018, Zero Waste International Alliance (ZWIA) was defined it as ‘the conservation of all resources through responsible production, consumption, reuse, and recovery of all products, packaging, and materials, without burning them and without discharge to land, water, or air, which may threaten the environment or human health.’ [19]. According to Hannon & Zaman [21], zero-waste is a catalyst that can encourage the participation of local communities to build sustainable cities for the future. Zero waste is considered a concept that goes beyond ‘generating no waste’ and is part of the resource recirculation society, which believes that waste is a resource.

Previous overseas studies related to zero waste were mainly related to industrial field and resource recirculation [21], recycling insurance and pre-recycling methods [22], and zero-waste cities [20, 23–24, 25], with a focus on building resource recirculation cities with cooperation between governments and industries. However, local research has been limited to passion industries, design, and resource utilization [26-29], and there has been no study related to health and healthcare.

To this end, the present study identified how the knowledge and attitude of zero-waste affected the behavior of students who spent relatively more time in their living spaces than other age groups because of online classes during the COVID-19 pandemic. Based on previous reports on the adverse effects of microplastics on human health, the present study aimed to lay a foundation for program development promoting zero-waste activities.

## Materials and Methods

### Study design

This was a cross-sectional study designed to identify how the knowledge and attitude of zero waste affected the behavior of students during the COVID-19 pandemic, when the usage of disposables was increased.

### Subjects and data collection

The study was conducted from August 20, 2021, to September 10, 2021, including university students in G metropolitan city. The convenience sampling method was used to assess subjects who agreed to the study purpose. The minimum sample size required for regression analysis was 173 subjects using the G Power 3.1 Program and considering the significant level of 0.5, power of 0.95, and total predictive factor of 14 in linear multiple regression analysis. Considering 20% dropout rate, the survey was conducted on 197 subjects, and the data of 196 subjects were analyzed.

Data were collected following the approval of the Institutional Bioethics Committee of E University (EU21-061). Online surveys were conducted after the study participants were informed of the purpose of the study, and they provided consent for data collection.

### Study tools

#### General characteristics

As general characteristics of the study subjects, age, sex, school year, major, and family type were assessed. The majors were categorized as health and medicine, natural science and engineering, education, humanity and social science, and others. The family types were classified as single-member households and two or more-member households.

#### Change in the usage of disposables after COVID-19 outbreak

To verify the change in the usage of disposables after COVID-19 outbreak, the following two questions were asked: ‘After COVID-19 outbreak, do you experience a change in the usage of delivery apps?’ and ‘After COVID-19 outbreak, do you experience a change in usage of parcel delivery service?’ two answers were provided, ‘no change’ and ‘increase’.

#### Zero-waste knowledge focusing on microplastics

Questions related to the knowledge of zero waste focusing on microplastics were developed by reviewing the literature and searching social networking sites (SNS), the Internet, and newspaper articles. The suitability of questions was verified by three professionals and five zero-waste executors.

Knowledge was divided into three categories: the generation process of microplastics, the health impact of microplastics, and environmental preservation. There were five questions regarding the generation process of microplastics: “I have never heard of microplastics,” “microplastics are generated during the disposal of plastic containers, there are microplastics in toothpaste and cosmetics, microplastics are reproduced by sunlight,” and “waste has come a full circle and come to my table.” There were six questions regarding the health impact of microplastics: “plastic itself contains carcinogens,” “microplastics lead to the accumulation of residual contaminants in the human body,” “microplastics cause systemic inflammation and immunosuppression,” “intake of microplastics causes cough, labored respiration, and pulmonary function insufficiency,” “microplastics can travel through blood vessels,” and “disposable cups contain substances causing an inflammatory response and adenocarcinoma.” There were four questions regarding environmental preservation: “I know what zero waste campaign is, I know environment-friendly enterprises, I know what is a recycle symbol,” and “I know some environmental policies, such as collecting empty bottles and tumbler discounts.”

Three answers were provided: “Yes, No, or Not sure.” “Yes” was assigned one point, and “No” and “Not sure” were assigned zero points. Points for each category were summed. The generation process of microplastics was scored from 0 to 5, and the environmental preservation and health impact of microplastics were scored from 0 to 4. Cronbach’s alpha for this tool was 0.745.

#### Zero-waste attitude

Questions related to zero-waste attitudes were developed by reviewing the literature and searching SNS, the Internet, and newspaper articles. Three professionals and five zero-waste executors verified the suitability of the questions.

The attitude was classified into five categories: eco-friendly production by companies, purchasing eco-friendly products, using eco-friendly products, separating disposables, and environmental campaigning. There were two questions regarding eco-friendly production by companies: “It is important to make products from materials that can be recycled” and “a company should make eco-friendly products”. There were four questions on purchasing eco-friendly products: “I think the things that I do not need are trash, I think the more eco-friendly products are, the better, carrying something like a tumbler is inconvenient, and It is important to use less disposable packaging.

There were three questions on using eco-friendly products: “It is boring to use purchased products for a long time,” “It is convenient to use straws, wooden chopsticks, and plastic bags,” and “It is convenient to use disposable wet wipes.” There were five questions on separating disposables: “eventually, it is beneficial for me to reduce the usage of disposable containers”, “the problem of disposable waste does not directly affect me”, “it is meaningless to make an effort to reduce the usage of disposables”, and “I feel uncomfortable generating plastic waste”. There were two questions on environmental campaigning: “I closely follow environmental campaigns” and “I have thought about participating in an environmental campaign”.

Each question was answered based on a 5-point Likert scale, with “strongly disagree” assigned one point and “strongly agree” assigned five points. Among the questions, negative responses for zero waste were processed as reverse questions. Average scores for each category are presented. A higher score indicated a positive attitude. Cronbach’s alpha for this tool was 0.731.

#### Zero-waste behaviors

Questions related to zero-waste behavior were developed by reviewing the literature and searching for SNS, the Internet, and newspaper articles. Three professionals and five zero-waste executors verified the suitability of questions.

Behavior was classified into four categories: purchasing eco-friendly products, using eco-friendly products, separating disposables, and environmental campaigns. There were five questions on purchasing eco-friendly products: “I check the recycle mark before buying something”, “I reduce waste by only purchasing what I need”, “I purchase products using as less disposable packaging as possible”, “I use eco-friendly products if it is possible”, and “if possible, I select no disposable check box when I order delivery food”. There were four questions on using eco-friendly products: “I keep using a product once I purchase it, I reuse daily necessities with containers by refilling them, I try not to use disposable wet wipes, and “I do not use disposables when I have reusable dishware”. There were four questions on separating and sending out disposables: “I actively separate and send out food and plastics, I try not to use delivery apps and parcel delivery services as much as possible because they generate much disposable waste, I remove the plastic packaging of PET bottles before taking them out to prevent generating mixed waste, and “I empty and clean recyclable plastic items before taking them out”. There were two questions on environmental campaigns: “I participate in empty bottle collection and tumbler discount” and “I reduce disposable waste by using reusable shopping bags”.

Each question was answered based on a 5-point Likert scale, with “strongly disagree” assigned one point and “strongly agree” assigned five points. Among the questions, negative responses for zero waste were processed as reverse questions. Average scores for each category are presented. Higher scores indicated pro-zero-waste behavior. Cronbach’s alpha for this tool was 0.767.

### Data analysis

Statistical analyses were conducted using SPSS (version 26.0; IBM SPSS Statistics, NYC, USA). The general characteristics and variables of the participants are presented as means and standard deviations or frequencies and percentages. The subjects’ general characteristics and differences in zero-waste behavior depending on the usage change of disposables after COVID-19 were analyzed using a t-test and ANOVA, respectively, followed by Scheffé’s post hoc test. Hierarchical multiple regression analysis was used to investigate the effect of zero-waste behavior, controlling for sex, age, school year, and major. In Model 1, the family type and usage change of disposables after the COVID-19 outbreak were entered. In Model 2, zero-waste knowledge was entered as a subcategory in Model 1. In Model 3, zero waste attitude was entered as a subcategory in Model 2.

## Results

### General characteristics of the study subjects

The general characteristics of the participants are presented in Table 1. A total of 196 participants, including 34 men (17.3%) and 162 women (82.7%), were included. The mean age of participants was 20.9 years. Regarding the school year, there were 48 (24.5%) first-year students, 33 (16.8%) second-year students, 79 (40.3%) third-year students, and 36 (18.3%) fourth- or higher-year students. Regarding major, 45 (35.2%) participants studied health and medicine, 36 (18.4%) studied natural science and engineering, 45 (23%) studied education, and 82 (39.3%) studied anthropology, sociology, and arts. Regarding family type, 25 participants (12.8%) lived alone and 171 (87.2%) lived with their families. Regarding the usage of disposables due to COVID-19, 136 (69.4%) participants reported increased usage. Regarding the usage of delivery apps due to COVID-19, 135 (68.9%) participants reported increased usage. Regarding the usage of parcel delivery services, 134 (68.4%) participants reported increased usage.

**Table 1.**
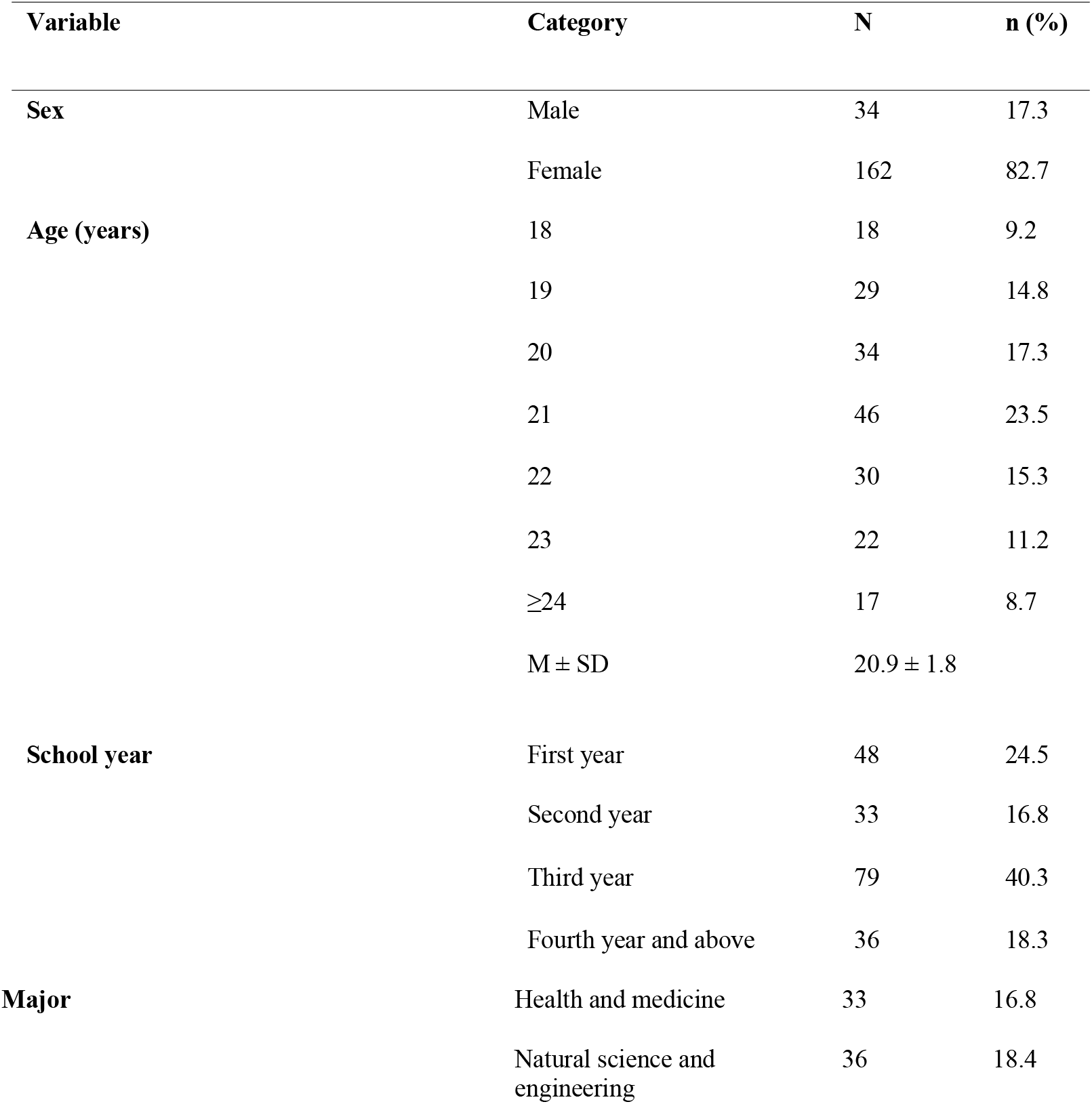
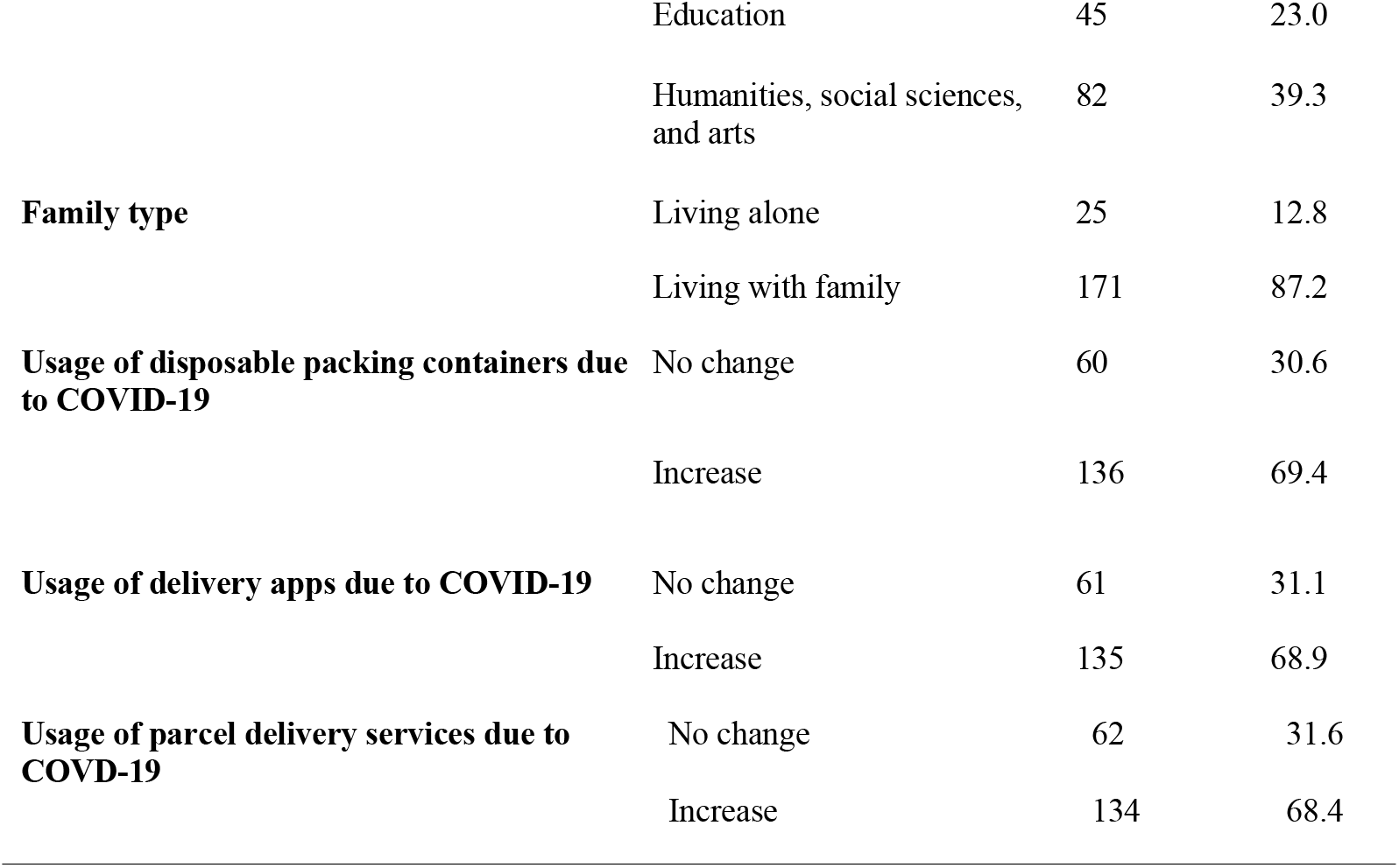
General characteristics of the subjects (n = 196)

| Variable | Category | N | n (%) |
| --- | --- | --- | --- |
| Sex | Male | 34 | 17.3 |
|  | Female | 162 | 82.7 |
| Age (years) | 18 | 18 | 9.2 |
|  | 19 | 29 | 14.8 |
|  | 20 | 34 | 17.3 |
|  | 21 | 46 | 23.5 |
|  | 22 | 30 | 15.3 |
|  | 23 | 22 | 11.2 |
|  | ≥24 | 17 | 8.7 |
|  | M ± SD | 20.9 ± 1.8 |  |
| School year | First year | 48 | 24.5 |
|  | Second year | 33 | 16.8 |
|  | Third year | 79 | 40.3 |
|  | Fourth year and above | 36 | 18.3 |
| Major | Health and medicine | 33 | 16.8 |
|  | Natural science and engineering | 36 | 18.4 |
|  | Education | 45 | 23.0 |
|  | Humanities, social sciences, and arts | 82 | 39.3 |
| **Family type** | Living alone | 25 | 12.8 |
|  | Living with family | 171 | 87.2 |
| **Usage of disposable packing containers due to COVID-19** | No change | 60 | 30.6 |
|  | Increase | 136 | 69.4 |
| **Usage of delivery apps due to COVID-19** | No change | 61 | 31.1 |
|  | Increase | 135 | 68.9 |
| **Usage of parcel delivery services due to COVID-19** | No change | 62 | 31.6 |
|  | Increase | 134 | 68.4 |

### Zero-waste knowledge, attitude, and behavior

Regarding zero-waste knowledge, the score for the MP generation process was 3.1/5 points and that for environmental preservation was 2.6/4 points. The score for the health effects of microplastics was 3.1/6. The average score for this category was 8.8/15.

Regarding zero-waste attitude, the score for eco-friendly production by companies was 4.4/5 points, that for purchasing eco-friendly products was 3.7/5 points, that for using eco-friendly products was 2.8/5 points, that for separating disposables was 4.1/5 points, and that for environmental campaigning was 3.6/5 points. The average score for each category was 3.7/5.

Regarding zero-waste behavior, the score for purchasing eco-friendly products was 3.5/5 points, that for using eco-friendly products was 3.7/5 points, that for separating disposables scored 4.0/5 points, and that for environmental campaigning was 3.6/5 points. The average score for the category was 3.7/5 points (Table 2).

**Table 2.**
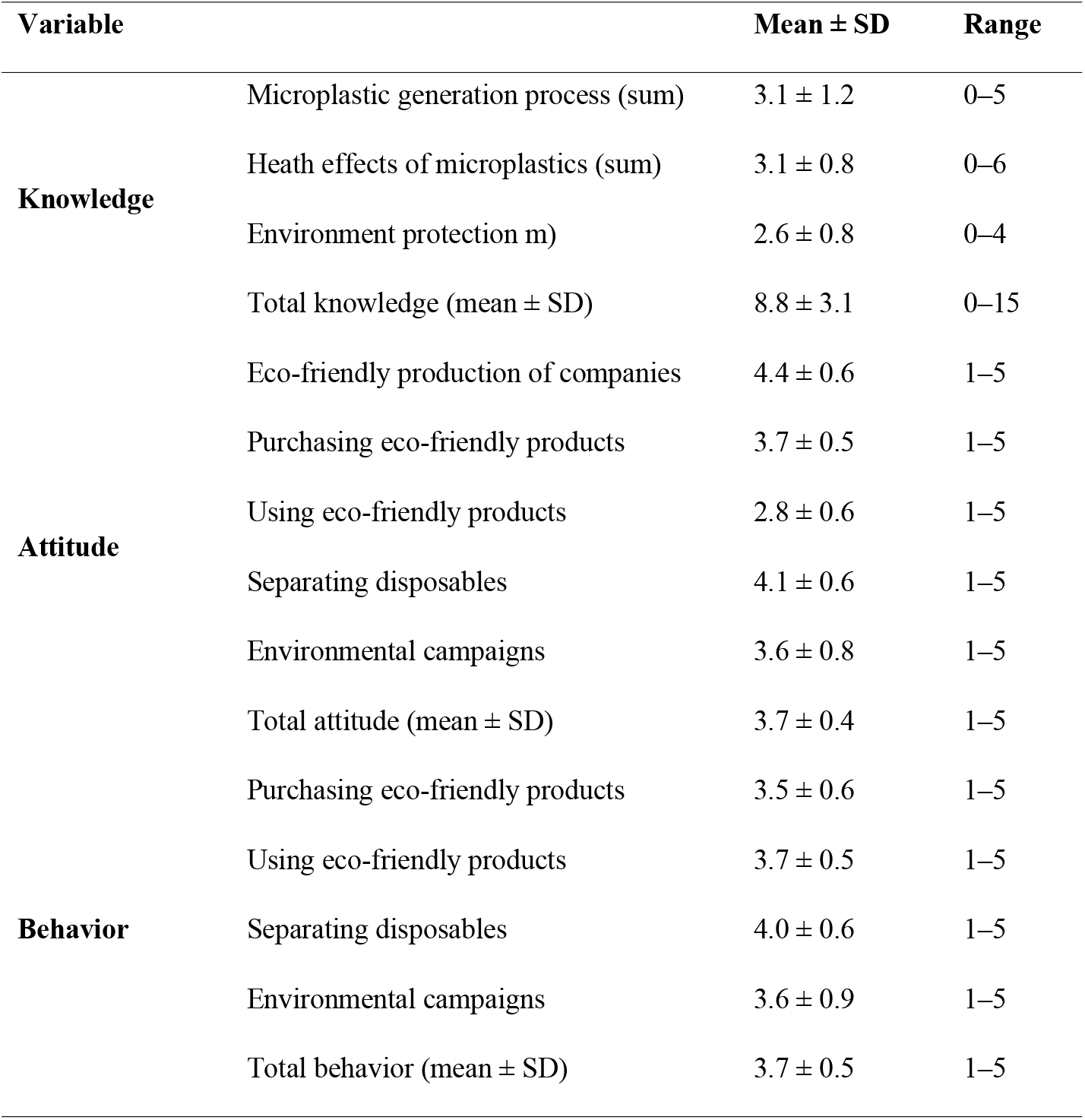
Zero-waste knowledge, attitude, and behavior (n = 196)

### General characteristics and differences in zero-waste behavior due to usage change of disposables after COVID-19 outbreak

Zero-waste behaviors, depending on the general characteristics of the study subjects, revealed significant differences by sex (t=-3.632, p=0.001) and family type (t=-2.324, p=0.021). Zero-waste behaviors showed significant differences in the usage of disposables after the COVID-19 outbreak (t=-2.454, p=0.015) (Table 3).

**Table 3.**
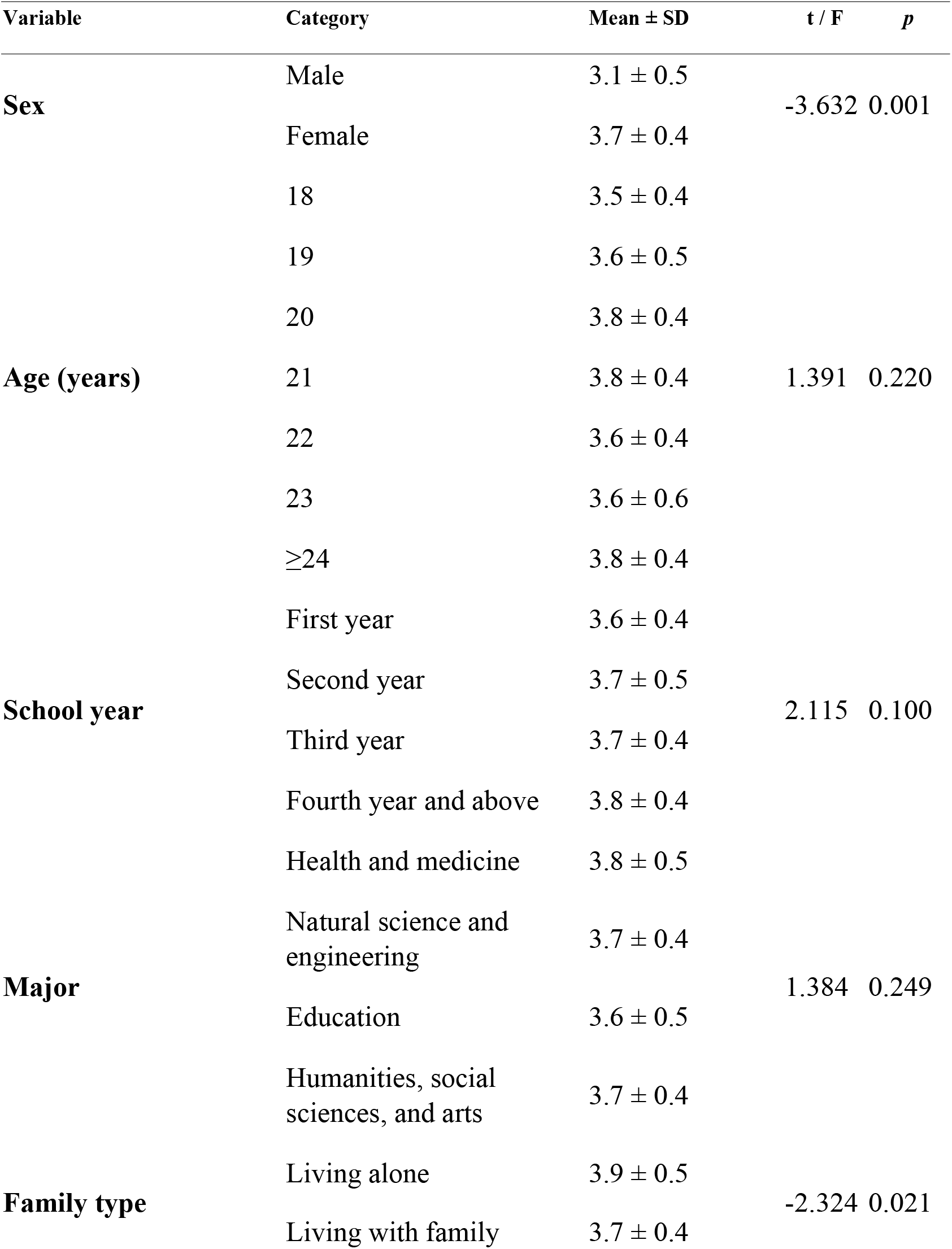
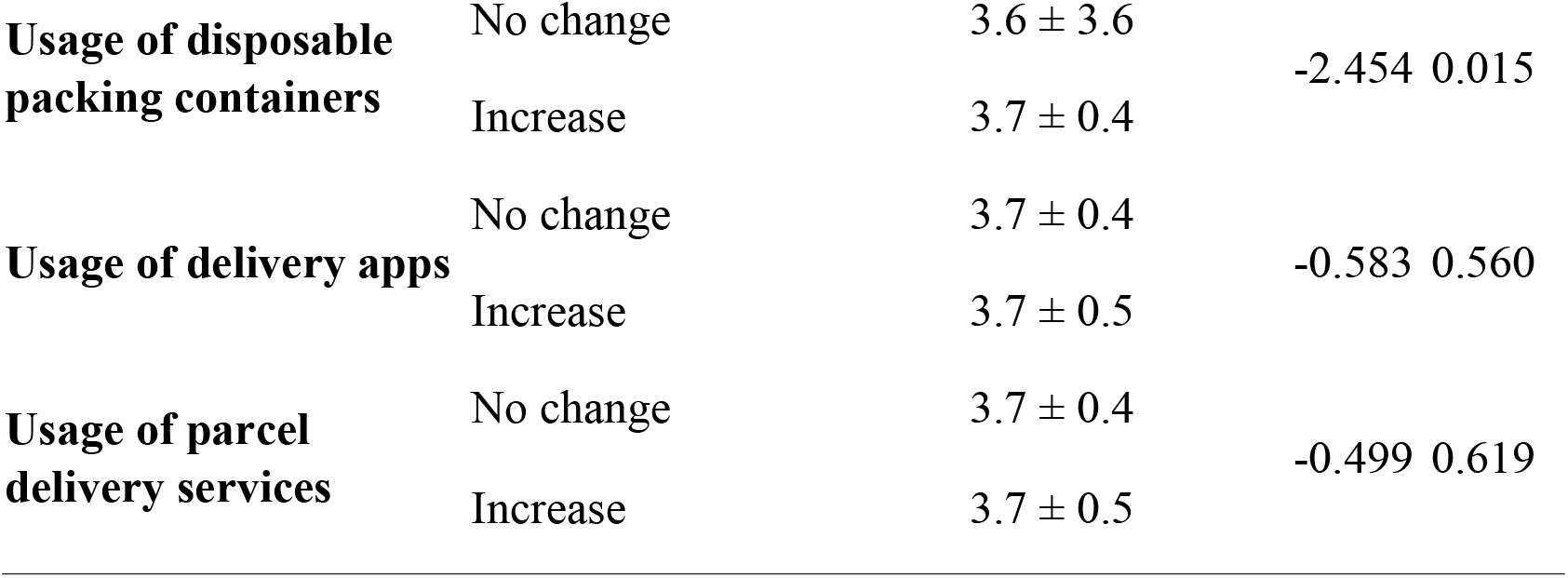
General characteristics and differences in zero-waste behaviors depending on the usage change of disposables after the outbreak of COVID-19 (n = 196)

### Factors influencing zero-waste behavior

Hierarchical multiple regression analysis was conducted to identify the factors affecting zero-waste behavior (Table 4). Sex, age, school year, and major were entered in each hierarchical regression model. The Durbin–Watson statistic was used to assess multicollinearity for verifying the basic assumption of regression analysis. The Durbin–Watson value was 2.232, and multicollinearity was low (variance inflation factor [VIF] < 5).

**Table 4.**
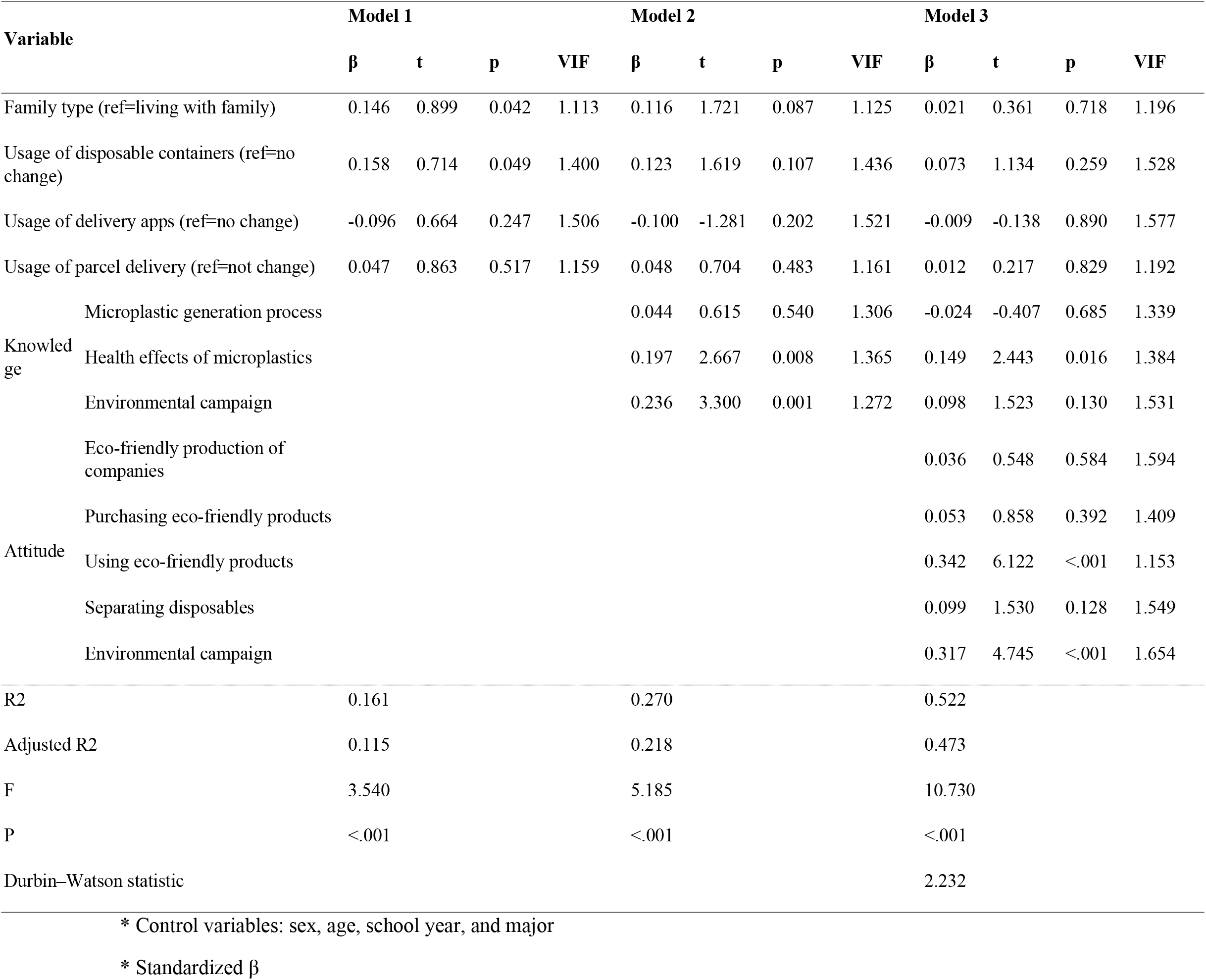
Factors influencing zero-waste behavior.

In Model 1, family type (β=0.146, p=0.042) and disposable usage (β=0.158, p=0.049) were significant factors. The regression model was significant (F=3.540, p<0.001) and the explanatory power was 11.5%.

Model 2 included the subcategories of zero-waste knowledge. The health effects of microplastics (β=0.197, p=0.008) and environmental campaign (β=0.236, p=0.001) were significant factors. The regression model was significant (F=5.185, p<0.001), and the explanation power was 21.8%.

In Model 3, the subcategories of zero-waste attitude were included. The health effects of microplastics (β=0.149, p=0.016), use of eco-friendly products (β=0.342, p<.001), and environmental campaign (β=0.317, p<0.001) were significant factors. The regression model was significant (F=10.730, p<0.001), and the explanation power was 47.3%.

## Discussion

The present study was undertaken to lay a foundation for program development aimed at improving zero-waste behavior by identifying the effects of zero-waste knowledge and attitude. Overall, during the COVID-19 pandemic, the usage of disposables amongst university students has increased.

In our analysis, score for the health effects of microplastics was the lowest, at 3.1/6 points, in the zero-waste knowledge category. Moreover, only the knowledge of health effects of microplastics affected zero-waste behavior amongst the knowledge categories. In other words, knowledge of the adverse link between microplastics and health was a major factor promoting zero-waste awareness and behavior. Previous studies have reported the detection of microplastics in table salt [13], drinking water [14], air [15], and marine organisms [11-12]. Various adverse effects of MPs on marine organisms have been reported [11-12]. Contamination of natural resources by microplastics enables their entry into the food chain and thus into the human body [29]. The accumulation of microplastics in human liver, kidney, and intestines disrupts energy and lipid metabolism [30]. However, the participants of the present study showed a low level of knowledge regarding the health effects of microplastics. Thus, health information related to microplastics should be provided. However, such education should be based on empirical studies of microplastics and its health risk. Animal experiments and studies have proven the risk of microplastics in other organisms [31-32]; however, no study has confirmed the health hazards in humans. This is because studies on the environment and its health effects are extensive, and the health risk appears over the long-term rather than the short-term. Furthermore, health risk differs amongst individual, rendering the identification of health problems caused by the environment difficult [33]. Studies collecting fundamental data for building scientific knowledge in the long-term are warranted to promote awareness regarding the health risks of microplastics.

In the category of zero-waste attitude, eco-friendly production of companies achieved the highest scores, whilst the usage of eco-friendly products achieved the lowest scores. The usage of eco-friendly products includes using the purchased products for a long time, convenience of using disposables, and convenience of cleaning with disposable wet wipes. According to a study on environmental problems by IPSOS [34], 91% of Korean respondents answered that there was concern regarding packaging waste and using disposables that cause environmental pollution. In terms of individual behavior to reduce unrecyclable packing materials, 27% of respondents answered that they tried to minimize their use by overcoming the habit of buying disposables, which was a low percentage, consistent with the results of the present study.

In another study, respondents answered that they occasionally use disposables because they are convenient and cheap. Nonetheless, with increased awareness of the environmental regulation policy, respondents tried avoiding the use of disposables to protect the environment [35]. In 2018, the Ministry of Environment announced a plan aimed at shifting the linear economic structure involving production and disposal to a circulating system involving production and recycling by 2027 [36]. Accordingly, regulatory measures were enacted to restrict the use of replaceable disposables and minimize unnecessary excessive packaging. However, this policy was modified and the use if disposables was allowed after the COVID-19 pandemic [37]. Accordingly, the usage of disposables increased due to COVID-19, and compliance with zero-waste behaviors became challenging as the non-use of disposables led to customer inconvenience. Therefore, more promotions and campaigns are required to encourage people to change their habits and inculcate zero-waste behaviors. Furthermore, companies should develop various alternative products that customers can use conveniently, such as tumblers, and offer them with a wide range of choices.

The present study showed that more interest in environmental campaigns and a positive attitude towards participating in such campaigns led to positive effects on zero-waste behaviors. Recently, Korea introduced environmental issues to the educational curriculum [38]. In elementary schools, education on environmental pollution is mandatory, although there is insufficient knowledge of environmental practices and participation in middle and high school curricula [39]. Furthermore, in the Korean education system, which focuses on college entrance examinations, it is difficult for students to acquire information on environmental issues and to have the opportunity to think about them by themselves. Therefore, students must be offered more opportunities to gain sufficient knowledge about environmental issues. This will help them acquire reliable information, promote zero-waste behaviors, and foster thinking on minimizing environmental pollution or microplastic usage. Additionally, various exciting public relations campaigns should be developed to encourage people.

Based on our results, the increased usage of disposables, delivery apps, and parcel delivery services due to COVID-19 did not affect zero-waste behavior. According to a previous study, the increased usage of delivery food due to COVID-19 has altered eating habits [40]. Students must use delivery apps and disposable containers to avoid contracting COVID-19; this increase is expected to be temporary during the COVID-19 pandemic. However, according to the Institute of Medicine, more infectious diseases are expected to emerge, indicating other possible outbreaks in the future [41]. Given the prolonged COVID-19 pandemic, awareness regarding the health effects of microplastics, importance of using recyclable products, and concerns for the environment should be promoted.

There were some limitations in the present study. First, the study subjects were limited to a particular age group rather than all age groups. University students are intelligent, and other factors may be added for different age groups. Second, although the measurement tool was developed with much effort, but it could not assess the comprehensive effect of microplastics and waste. Therefore, additional tools should be created. Finally, this was a cross-sectional study undertaken at a specific time when the COVID-19 pandemic extended. Additional studies on zero-waste behaviors in the long term are warranted. Despite these limitations, our study identified factors affecting zero-waste behavior and contributed to environmental research. Zero-waste behaviors may vary depending on age and presence/absence of chronic diseases. Future studies should include other subject groups. In recent years, the infertility rate has been rising, while the birth rate has reduced. Therefore, additional studies are required to verify the link between reproductive health and microplastics in the environment.

## Data Availability

all contained within the manuscript

## Data Availability Statement

All relevant data are within the paper.

## Funding statement

The funders had no role in study design, data collection and analysis, decision to publish, or preparation of the manuscript.

## Authors contribution

Eun-Hi Choi: Conceptualization, Data curation, Formal analysis, Methodology, Supervision, Validation, Visualization, Writing – original draft

Hyunjin Lee: Supervision, Validation, Writing – original draft, Writing – review & editing

Mi-Jung Kang: Supervision, Validation, Writing – original draft, Writing – review & editing

Inwoo Nam: Conceptualization, Methodology, Data curation, Investigation

Hui-Kyeong Moon: Conceptualization, Methodology, Investigation

Ji-Won Sung: Conceptualization, Methodology, Investigation, ‡ IN, HKM and JWS also contributed equally to this work.

Jae-Yun Eu: Data curation, Methodology, Investigation

Hae-Bin Lee: Data curation, Methodology, Investigation

## Notes

### Competing Interest Statement

The authors have declared no competing interest.

### Funding Statement

The authors received no specific funding for this work

### Author Declarations

the Institutional Bioethics Committee of Eulji University (EU21-061)

